# Identifying adverse experiences in childhood in the Born in Bradford Birth Cohort children

**DOI:** 10.64898/2026.08.18.26360723

**Authors:** Natalie Lam, Ruth Wadman, Aidan Watmuff, Simon Gilbody

## Abstract

Adverse experiences in childhood (AEs) typically refer to undesirable events, including child maltreatment and household challenges. Various survey measures and linked routine data in the Born in Bradford Birth Cohort (BiB) datasets can provide a contemporary understanding of the distribution of AEs in the population and the factors related to their occurrence. This study aimed to identify relevant survey data on AEs collected from BiB families and to summarise the prevalence of AEs from birth to early adolescence (ages 12-15) among BiB children. We included BiB children who participated in the follow-ups – Growing Up (GUp, n=5253) and Age of Wonder (AoW, n=2662). Four AEs were identified – parental mental illness, parental substance use, children not living with both parents in the same home, and being bullied by peers. The survey data included 1) health, substance use, living arrangements, and children’s bullying experience reported by parent(s) at baseline (2007-2011, around birth) and/or GUp (2017-2022, during mid-childhood), and 2) bullying experience and living arrangements self-reported by children at AoW (2022-2024, during early adolescence). Additionally, we included parents’ primary care records regarding any mental illness or substance use. Overall, 3371 (64.2%) children experienced at least one of the four AEs between birth and early adolescence. The most common AE was parental mental illness, whereas parental substance use was the least common. Children across all sociodemographic groups experienced AEs. Asian children, or those whose mothers were not materially deprived, appeared less likely to experience AEs. Conversely, children of White or Mixed ethnicities, or whose mothers were materially deprived, were more likely to experience AEs. Consistent with similar studies, our findings show that AEs are widespread but disproportionately affect certain sociodemographic subgroups among BiB children. These disparities can be reduced by early-years policies that provide practical family support, guided by continuously collected AE data.

## Introduction

Adverse experiences in childhood, or childhood adversities, hereafter referred to as “AEs”, usually denote undesirable events or a series of child maltreatment incidents (i.e. abuse and neglect) and household challenges (e.g., problematic substance use within the household). These data are not easily collected locally or nationally, particularly during childhood [1, 2]. The fact that existing studies used various survey questionnaires and routine records to estimate the prevalence of AEs in a population or study sample indicates that no single, consistent information source documents the occurrence of a range of AEs across the child population’s childhood.

AEs have been recognised as a public health concern due to how common they are in the UK population and their links to long-term negative impacts on individuals’ physical and mental health and life chances [3–5]. Children from all socioeconomic backgrounds experience AEs. However, many studies in the UK and globally have found that those from more disadvantaged or deprived socioeconomic backgrounds are more likely to encounter AEs [1, 6]. AEs are of particular concern for the communities and professionals who engage with children and families in Bradford, which is one of the most deprived cities [7], with a high proportion of child population in the UK [8, 9]. Moreover, Bradford has the second-highest proportion of children living in relatively low-income families [9]. Considering the higher proportions of the child population and children under 16 living in low-income families, a greater number and proportion of children in Bradford are believed to have experienced AEs compared to other areas in the UK [2, 7]. The health needs assessment of adverse childhood experiences (ACEs) in Bradford reported that some of the AEs, e.g., parental mental health problems and parental substance use, were higher than the national (within England) and regional (within Yorkshire and Humber) averages [2]. However, it also revealed that accurate AE prevalence data were unavailable at the local level, particularly regarding parental separation and parental incarceration, despite access to routine records from health and social care and police data.

Various measures in the Born in Bradford Birth Cohort (BiB) datasets, including surveys and data linkage with routine data, can provide a contemporary understanding of the distribution of (at least some) AEs in the population and the risk factors associated with their occurrence. Combining data from different information sources, e.g., surveys and routine records, can supplement the gaps in each source to provide a more comprehensive estimate of AE prevalence. This method is not uncommon in AE studies, e.g., in studies by Fagan *et al.* [10] and Morrow *et al.* [11]. In the Born In Bradford Birth Cohort (BiB) Study of over 13,000 children born in 2007-2011, data linkage to the routine records of child and parent participants has been established. Additionally, longitudinal data about the children and their families are collected via surveys from the mother’s pregnancy across the child’s life course. The details include socioeconomic characteristics, demographic information (e.g., household size, living arrangements), lifestyle (e.g., substance use), environment and neighbourhood, physical and mental health of both parents and children, and the development and experiences of the children (e.g., being bullied). Many of these details are pertinent to the factors influencing the occurrence and exposure to AEs among BiB children.

### Aims

This study aimed to identify BiB survey data relevant to AEs experienced by children born and growing up in Bradford, and to summarise the sample prevalence of AEs among BiB children from birth to early adolescence (ages 12-15). It also explored the feasibility of using existing linked routine healthcare data to supplement the survey data where suitable.

## Methods

We followed the Strengthening the Reporting of Observational Studies in Epidemiology (STROBE) Statement [12] (see checklist in S1 Supporting Information). This study presents descriptive statistics from a longitudinal study, which analysis plan was pre-registered on OSF before the data analysis began [13].

### Criteria for adverse experiences in childhood

To identify the data variables relevant to AEs in the BiB datasets, we established a list of AE criteria for classifying AEs. This set of criteria is shaped around a child and two of a child’s immediate environments, namely home and school, concerning safeguarding and promoting the welfare of the child, whilst adopting the definitions from Kalmakis and colleagues [14] and Kelly-Irving and colleagues [15]. An AE is/does:

- An event or a series of events, or actions that happen during childhood, i.e. from birth to before 18 years of age, directly influences the child, or is directly inflicted upon or witnessed by the child, in the context of relationships or personal interaction between the child and others;
- Happen within the child’s family or school, which are two of their immediate and regular environments (micro- and meso-systems);
- Vary in timing and frequency of occurrence, and self-perceived severity;
- Lead to an elevated risk of the child experiencing harm and distress, and detrimental impacts on physical, emotional, and educational needs; thus, health and social outcomes across the life course; and
- Typically, include the notions and actions of maltreatment, harm, and unpleasant or disadvantageous deviation from societal norms, where possible to be distinguished from conditions in the socioeconomic and material environment which are excluded from this project.

Among the categories commonly referred to as AEs in children’s services, policies, and literature, eleven categories of AEs meet these criteria (see **Error! Reference source not found.**). The first ten categories were the adverse childhood experiences (ACEs) measured in the CDC-Kaiser ACE Study [16, 17]. Peer victimisation, which includes bullying, is the eleventh category. Perceptions of the occurrence and severity of these experiences vary between individuals and societies. Nevertheless, the above criteria are generic and likely to be widely applicable across cultures and societies.

### AE classification process overview

To achieve this study’s aims, we adhered to the five principles set below to decide which information sources and variables to use for classifying AEs. These principles were informed by the childhood adversity literature and a scoping review [18] on common methods for measuring and classifying AEs. The five principles are:

1. Focusing primarily on survey datasets as the main source for identifying AEs for classification, then supplementing with GP records where relevant and feasible. That is, when an AE can be identified from survey data, any available relevant GP records are also used to construct the overall AE category.
2. A classified AE matches one of the eleven AEs of interest listed in **Error! Reference source not found.**, as well as the corresponding questions and their meanings used to measure that AE in the included studies identified in the cited scoping review [18].
3. AE occurrence is considered binary and classified as “yes” or “no” regarding its presence at a time point or over a period.
4. To establish the temporality of the AE occurrence if possible, i.e. timing and duration, or at least referring the presence to the measurement time point.
5. Where the presence of an AE is identified from parental data, such as parental mental illness, the child is classified as having experienced that AE, regardless of whether the parents lived with the child.

These five principles were operationalised into the following four main tasks for the classification of AEs:

1. To identify a combination of BiB surveys, from two or more time points, while maximising the number of BiB children and families included in this study and ensuring the included sample is the same for each classified AE category;
2. To identify the survey data variables, reported by parents and/or children, that are relevant to the AEs of interest and can indicate their occurrence;
3. To select the Read Codes Clinical Terms Version 3 (CTV3) diagnostic codes in order to search the electronic GP records linked to the BiB datasets of eligible, consented participants. The reason for not using other healthcare records, e.g., secondary care, was that GP records usually contain a person’s broader healthcare information, including referrals and hospital admissions. Other routine data, specifically health visiting and social services records, which may contain indicators of other AEs (e.g., abuse and neglect) were not available for request via data linkage at the time of obtaining the data sharing agreement in January 2024 [19];
4. To combine the selected data variables to construct each corresponding AE category.

### Included BiB studies and participants

The classification of AE categories was primarily data-driven, while also attempting to align with common AE definitions in the literature and with children’s services practices and policies. While reviewing all variables in the BiB studies to identify those potentially relevant to the 11 AEs of interest, we identified survey variables that were both relevant and sufficient to classify children as having experienced bullying, parental substance use, parental mental illness, or not living with both parents between birth and early adolescence, using data from three of the largest BiB surveys highlighted in Table 2. Moreover, using these three surveys allows us to maximise the number of BiB children and families included in this study.

**Table 1.** Eleven adverse experiences in childhood of interest.

| Eleven AEs of interest |  |
| --- | --- |
| 1 | Emotional abuse by parental/caregiver |
| 2 | Physical abuse by parental/caregiver |
| 3 | Sexual abuse and exploitation |
| 4 | Emotional neglect by parental/caregiver |
| 5 | Physical neglect by parental/caregiver |
| 6 | Domestic abuse (parental/caregiver being treated violently or by coercive control within the home by his/her partner) |
| 7 | Parental/caregiver's substance abuse |
| 8 | Parental/caregiver's mental illness |
| 9 | Parental separation |
| 10 | Parental/caregiver incarcerated (or prosecuted) |
| 11 | Peer victimisation (experienced bullying, assault, physical intimidation, or emotional victimisation by a non-sibling peer) |

**Table 2.** Measurable versus unmeasurable AEs of interest within the four largest BiB surveys.

|  |  | Around birth | Mid-childhood |  | Early adolescence |
| --- | --- | --- | --- | --- | --- |
|  |  | Baseline | Primary School | GUp | AoW |
| 1 | Physical abuse | ✗ | ✗ | ✗ | ✗ |
| 2 | Emotional abuse | ✗ | ✗ | ✗ | ✗ |
| 3 | Sexual abuse and exploitation | ✗ | ✗ | ✗ | ✗ |
| 4 | Physical neglect | ✗ | ✗ | ✗ | ✗ |
| 5 | Emotional neglect | ✗ | ✗ | ✗ | ✗ |
| 6 | Parental/caregiver's mental illness | ✓ | ✗ | ✓ | ✗ |
| 7 | Domestic abuse (Family member being treated violently or by coercion control within the home) | ✗ | ✗ | ✗ | ✗ |
| 8 | Parental separation (or separation from parent(s) including death of parent(s) and being placed in care) | ✓ | ✗ | ✓ | ✓ |
| 9 | Incarcerated household member (or prosecuted) | ✗ | ✗ | ✗ | ✗ |
| 10 | Parental/caregiver's substance use | ✓ | ✗ | ✓ | ✗ |
| 11 | Peer victimisation (bullying, assault, physical intimidation, or emotional victimisation by a non-sibling peer) | ✗ | ✓ | ✓ | ✓ |

Notes: ✓ = measurable and can be classified or inferred; ✗ = not measurable and cannot be confidently classified or inferred; AoW = Age of Wonder survey; GUp = Growing Up survey; Green-shaded cells highlight the time points and exposure variables selected for this project.

Baseline BiB data were collected between 2007 and 2011, typically around 26-28 weeks of gestation, when the mothers consented to participation and data linkage, and completed the self-report questionnaire [20, 21]. The Growing Up Study survey (GUp) took place between March 2017 and January 2022 during mid-childhood [22, 23]. Regarding the Age of Wonder Study surveys (AoW), only data from the academic years 2022-23 and 2023-24 were available when data were requested for this BiB study in February 2025, and were therefore included. These surveys were conducted at three time points from baseline to early adolescence.

To summarise the eligibility criteria, out of all 13,858 BiB children, 5253 BiB children were included for the period between birth and GUp because their natural parent(s) participated in GUp. 2662 children were included for the period across baseline, GUp, and AoW, because their natural parent(s) participated in the GUp and they were recruited into the AoW during 2022-23 and/or 2023-24 academic years through schools’ participation.

The sample in GUp largely determined the overall sample sizes for children and parents, as it linked eligible participants at baseline with those at AoW. Of the 5253 included BiB children, 4632 mothers and 301 fathers (4933 parents; mother-to-father ratio = 16:1) completed the GUp survey. 302 children had both parents complete the GUp survey, while 4931 children had such data from mothers only, and 20 children had such data from fathers only. A total of 6049 parents (4651 mothers and 1398 fathers; ratio = 3:1) of these 5253 children consented to primary care data linkage at baseline, so their GP records were included in this study (see Figure 1).

**Figure 1.**
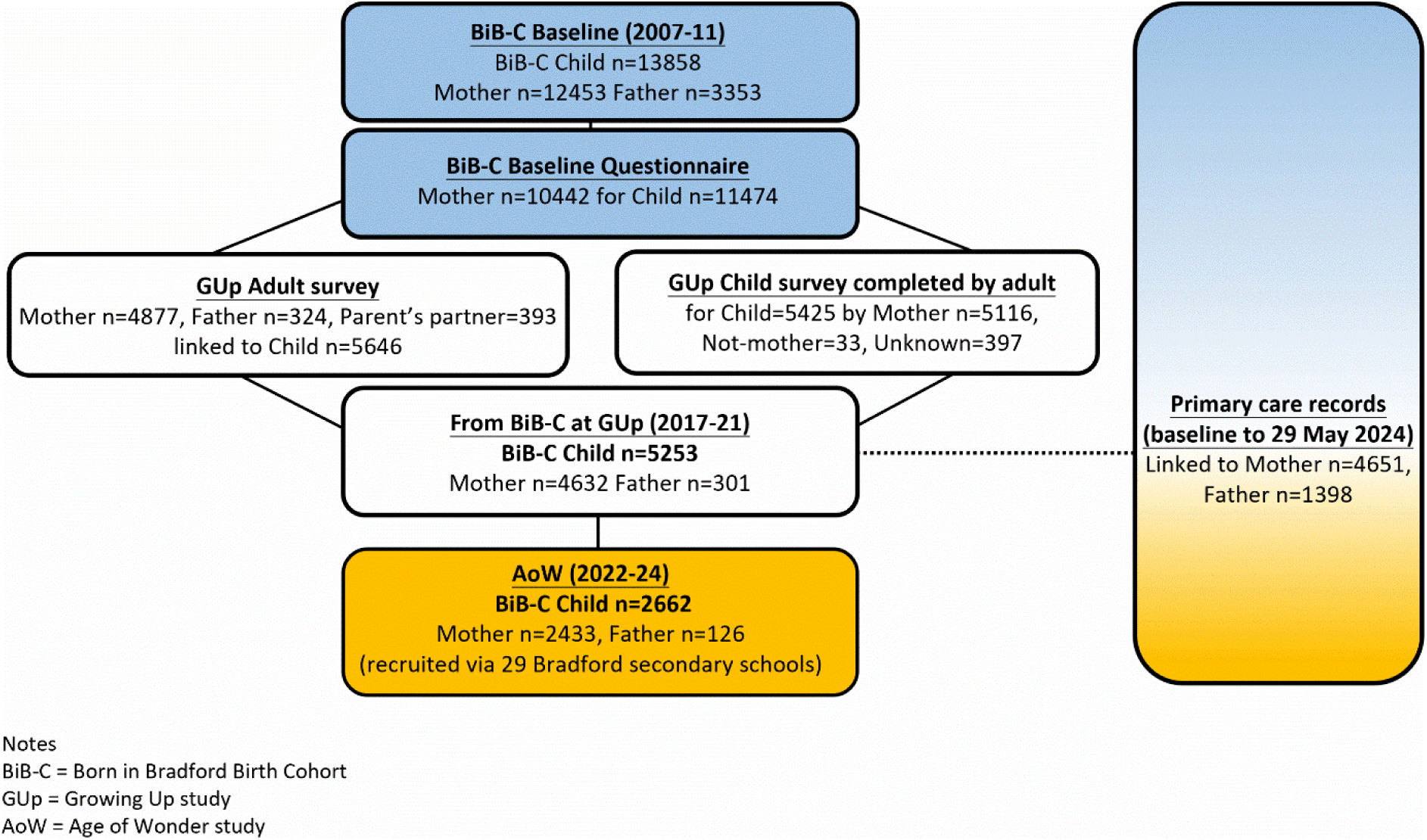
Born in Bradford Participants flow chart: data available for analysis, from baseline to Age of Wonder 2022-24.

### Identify the AE variables data in BiB surveys and GP records

Figure 2 summarises the data available in the surveys and GP records collected between baseline and AoW. When considering the temporality of each AE, we could confidently infer that the AEs were present between the time points, or shortly before or at a specific time point. However, it was not feasible to confirm the duration of occurrence of any of these AEs, or the exact timing of emergence of parental mental illness, parental substance use, or not living with both parents. As a result, the temporality and classification of each of these AEs are:

1. Being bullied: 1) at mid-childhood, and 2) at early adolescence;
2. Not living with both parents: 1) around birth, 2) at mid-childhood, and 3) at early adolescence;
3. Parental mental illness: at some point between 1) around birth and mid-childhood, and 2) around birth and early adolescence;
4. Parental substance use: at some point between 1) around birth and mid-childhood, and 2) around birth and early adolescence.

**Figure 2.**
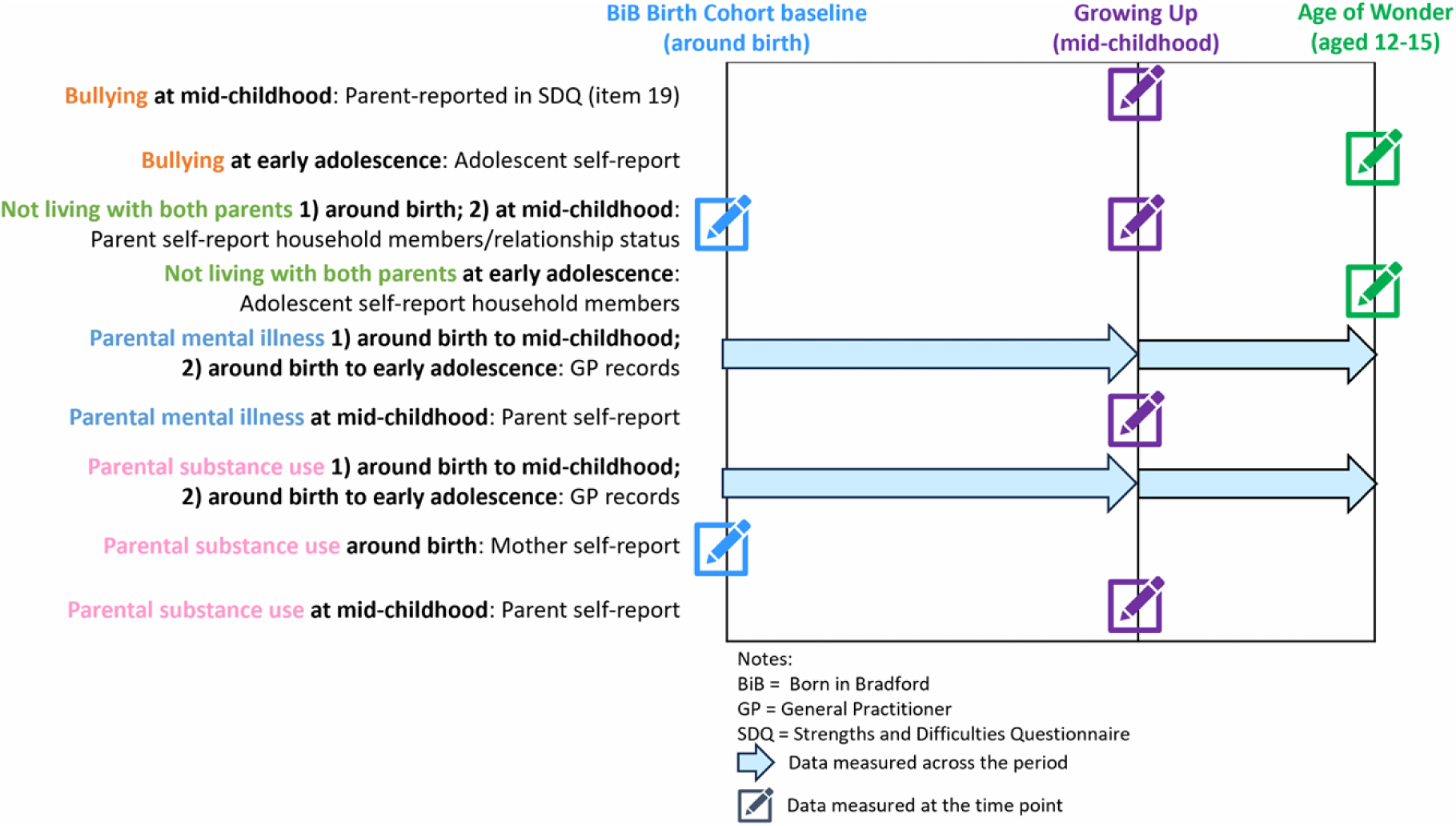
Data collection methods and time points for classifying adverse experiences in childhood.

There were no survey data which could be used to confidently infer seven of the AE categories of interest, which are physical abuse, emotional abuse, sexual abuse and exploitation, physical neglect, emotional neglect, domestic abuse, and incarcerated/prosecuted household member. The BiB survey variables used for AE classification are listed in S2 Supporting Information.

Survey data were the primary information source, while we used GP records as a supporting source. The rationale and methods for classifying each AE category are explained below. We first describe how we used the GP records, then how we constructed the four AE categories based on the available survey data and the identified GP records data.

### Selecting diagnostic codes for searching GP records

Clinical data routinely recorded by GPs were used to identify parental mental illness and substance use. GP records typically contain information about a person’s diagnosed conditions, appointments at the GP practice, prescriptions, and letters from hospitals or specialists sent to the GP [24]. The goal of using the parents’ GP record data was to identify any record of a diagnostic code related to mental illness or substance use during the childhood of their BiB children. At the time of data request, CTV3 codes were the set of clinical codes recorded in the BiB datasets. All CTV3 codes used are listed in S2 Supporting Information.

### Mental illnesses records

The mental illnesses included in this study were anxiety, depression, comorbidity of anxiety and depression, psychosis, schizophrenia, bipolar affective disease, and eating disorders. The phenotypes of eating disorders include six conditions listed in the ICD-10, namely anorexia nervosa, atypical anorexia nervosa, bulimia nervosa, atypical bulimia nervosa, overeating associated with other psychological disturbances, and vomiting associated with other psychological disturbances [25]. 432 CTV3 codes specifically denoted to these mental illnesses were identified through the process described in Appendix 1 in S3 Supporting Information. The included codes were based on the list of common and severe mental illness codes used and provided by Prady and colleagues [26].

### Substance use records

For substance use, 303 diagnostic codes relating to substance dependence, either alcohol or illicit drugs, were included. Only the CTV3 diagnostic codes which refer to ongoing or recent substance use conditions were identified for the classification. That is, the condition and concerns were likely present on the date of record or shortly before. The steps and information used to identify these codes are detailed in Appendix 2 in S3 Supporting Information.

A Data Analyst with access to the BiB data warehouse extracted the earliest recorded date of the first diagnostic code in the GP record of the consented parents, between their baseline dates and 29 May 2024. The results were then combined with other data for each participating parent and their participating child by using the unique participant identifier to match individuals and construct the parental mental illness and parental substance use categories.

### Constructing the four AE categories

The rationale and methods for classifying each AE category are detailed below, including how survey data and GP records were combined to form the four AE categories.

### Parental mental illness reported in surveys and from GP records

In GUp, parents self-reported any history of mental illness and clinically significant symptoms of depression on the Patient Health Questionnaire (PHQ-8) [27] or anxiety on the Generalized Anxiety Disorder 7-item (GAD-7) questionnaire [28] at the time of the survey. The method for classifying these survey data is detailed in Appendix 3 in S3 Supporting Information. From the GP records and GUp survey, we identified parents who self-reported having mental illness or had GP-recorded mental illness to infer their children experienced parental mental illness.

### Parental substance use reported in surveys and from GP records

Parental substance use typically refers to whether any parent consumes alcohol and/or illicit drugs to the extent that they become dependent on the substances, or their daily lives, health, and well-being are adversely affected by this consumption. Using parents’ survey data, we identified parents who self-reported illicit drug use and/or a hazardous pattern of alcohol consumption to infer that their children experienced parental substance use. Mothers’ drug use during pregnancy was surveyed at baseline, and parents’ alcohol consumption was measured in the baseline and GUp surveys. The method used to classify parental substance use is detailed in Appendix 4 in S3 Supporting Information.

### Children not living with mother or father reported in surveys

This AE category aimed to identify the children who did not live with both natural parents in the same home, daily or most of the time, at some point during their childhood. Although the reasons were not directly captured in the BiB datasets, parental separation, death of a parent, or a parent being away for an extended period, e.g., living abroad or incarcerated, could be some of the reasons. If a child did not live with any parent but with a guardian or foster carer, the child was also regarded as not living with both parents. Some of these situations adhere to the notions of commonly regarded ACEs, such as parental separation and parental incarceration. Additionally, this category may include other adverse experiences proposed by some researchers, e.g., parent deceased or child in care [1, 6]. Different sets of questions were used to collect living arrangement and household composition information at baseline, GUp, and AoW. The details used to classify that a child was not living with both parents at each of these time points are provided in Appendix 5 in S3 Supporting Information.

### Bullying reported in surveys

The bullying experience of a child refers to the child being bullied at some point or over time. More specifically, being bullied refers to being assaulted, physically intimidated, or emotionally victimised by one or more non-siblings. This is similar to peer victimisation, defined by Finkelhor *et al.* [29]. We used the parent-reported information in the Strengths and Difficulties Questionnaire (SDQ) [30] in GUp to indicate whether the child was bullied or not at mid-childhood. In the AoW surveys, adolescents were asked whether they were bullied in the previous two months, and self-rated on the SDQ in Year 9. The detailed methods for classifying bullying experiences are listed in Appendix 6 in S3 Supporting Information.

### Combining parents’ data

To classify parental substance use, parental mental illness, and not living with both parents, the available and relevant data regarding either parent were used. If only one parent’s data were available, i.e. the main respondent’s, the child was classified according to that parent’s data only. This classification approach is similar to other ACEs studies which use data from other birth cohorts, e.g., reported by Russell *et al.* [31] and Straatmann *et al.* [32]. Some children had both parents participated in GUp and/or consented to data linkage. If one or two parents of these children had substance use or mental illness, the children were classified as having experienced the corresponding AE.

### Data analysis

Descriptive statistics were conducted in R (Version 4.5.0) in RStudio (Version 2024.12.1+563 “Kousa Dogwood” Release (27771613, 2025-02-02) for Windows). The R packages used were *arsenal* (v3.6.3) [33], *compareGroups* (v4.9.1) [34], *dplyr* (v1.1.4) [35], and *tidyverse* (v2.0.0) [36]. The tables for descriptive statistics were produced using the *arsenal* package and its simulation function to simulate 5000 replicates for p-values for categorical variables against the levels of the stratifying variables. The tables were then copied and pasted into Microsoft Word and Microsoft Excel for Microsoft 365 MSO (Version 2602 Build 16.0.19725.20126) to format and produce the tables and graphs. The R code for constructing the AEs and conducting the analyses is available on GitHub, at https://github.com/NatalieYELam/adverse_experiences-/blob/main/AE-classification_Rcodes.html. To compare the number of reported cases of parental substance use and parental mental illness between information sources, we used 2×2 contingency tables.

All eligible participants were included in the data analyses. However, the details presented are the observed data only. There were approximately 1.9% to 16.4% missing data in each AE category between birth and mid-childhood, and 2.1% to 49.2% missing data in the AE categories between birth and early adolescence (see Table 3 Characteristics of the included children and families, stratified by surveysTable 3). Data were missing mainly due to various reasons of non-participation in surveys [21, 22, 37]. At baseline and GUp surveys, a higher proportion of missing data was related to ethnicity and maternal socioeconomic position. At AoW, the main reasons appeared to be primarily related to schools’ participation.

**Table 3.** Characteristics of the included children and families, stratified by surveys.

|  | BiB@GUp | BiB@GUp+AoW |  |
| --- | --- | --- | --- |
|  | Eligible children | Recruited to AoW | Not recruited to AoW |
|  | n=5253 | n=2662 | n=2591 |
| **Child's sex at birth** |  |  |  |
| Female | 2548 (48.5%) | 1392 (52.3%) | 1156 (44.6%) |
| Male | 2705 (51.5%) | 1270 (47.7%) | 1435 (55.4%) |
| **Has congenital anomalies (GP record linkage up to age 5)** |  |  |  |
| No Record | 4948 (94.2%) | 2522 (94.7%) | 2426 (93.6%) |
| Yes | 305 (5.8%) | 140 (5.3%) | 165 (6.4%) |
| **Child's ethnicity** |  |  |  |
| White | 1510 (28.7%) | 565 (21.2%) | 945 (36.5%) |
| Mixed | 213 (4.1%) | 79 (3.0%) | 134 (5.2%) |
| Asian | 3388 (64.5%) | 1957 (73.5%) | 1431 (55.2%) |
| Black | 65 (1.2%) | 23 (0.9%) | 42 (1.6%) |
| Other | 74 (1.4%) | 38 (1.4%) | 36 (1.4%) |
| Missing | 3 (0.06%) | 0 (0.0%) | 3 (0.1%) |
| **Age GUp survey taken<sup>1</sup>** | 9.72 (1.14) | 9.55 (1.10) | 9.89 (1.15) |
| **Age first AoW survey taken<sup>1</sup>** | -- | 13.8 (0.82) | -- |
| **IMD for address at BiB recruitment (2010 score)** | 43.3 (17.2)* | 45.2 (16.0)* | 41.4 (18.1)* |
| **Maternal socioeconomic position (birth)<sup>2</sup>** |  |  |  |
| Least dep | 884 (16.8%) | 380 (14.3%) | 504 (19.5%) |
| Employed not mat dep | 786 (15.0%) | 359 (13.5%) | 427 (16.5%) |
| Employed no money | 701 (13.3%) | 364 (13.7%) | 337 (13.0%) |
| Benefits but coping | 1411 (26.9%) | 777 (29.2%) | 634 (24.5%) |
| Most dep | 598 (11.4%) | 328 (12.3%) | 270 (10.4%) |
| Missing | 873 (16.6%) | 454 (17.1%) | 419 (16.2%) |
| **Number of people in mother's household (around birth)<sup>1</sup>** | 4.45 (2.41)* | 4.66 (2.46)* | 4.24 (2.34)* |
| **Mother's age at child's date of birth (years)<sup>1</sup>** | 28.1 (5.50) | 28.1 (5.36) | 28.1 (5.63)* |
| **Father's age at child's date of birth (years)<sup>1</sup>** | 31.1 (6.64)* | 30.9 (6.56)* | 31.2 (6.71) * |
| **Any AE (birth to GUp):** |  |  |  |
| Had AE | 3147 (59.9%) | 1535 (57.7%) | 1612 (62.2%) |
| No AE | 1586 (30.2%) | 860 (32.3%) | 726 (28.0%) |
| Missing | 520 (9.9%) | 267 (10.0%) | 253 (9.8%) |
| **Any AE (birth to AoW):** |  |  |  |
| Had AE | 3371 (64.2%) | 1747 (65.6%) | 1624 (62.7%) |
| No AE | 308 (5.86%) | 308 (11.6%) | 0 (0.0%) |
| Missing | 1574 (30.0%) | 607 (22.8%) | 967 (37.3%) |
| **Not lived with mother or father sometime (birth to GUp)** |  |  |  |
| Not both parents | 1203 (22.9%) | 537 (20.2%) | 666 (25.7%) |
| Both parents | 3108 (59.2%) | 1636 (61.5%) | 1472 (56.8%) |
| Missing | 942 (17.9%) | 489 (18.4%) | 453 (17.5%) |
| **Not lived with mother or father sometime (birth to AoW)** |  |  |  |
| Not both parents | -- | 681 (25.6%) | -- |
| Both parents | -- | 1055 (39.6%) | -- |
| Missing | -- | 926 (34.8%) | -- |
| **Lived with both parents around birth** |  |  |  |
| Not both parents | 536 (10.2%) | 244 (9.2%) | 292 (11.3%) |
| Both parents | 3857 (73.4%) | 1970 (74.0%) | 1887 (72.8%) |
| Missing | 860 (16.4%) | 448 (16.8%) | 412 (15.9%) |
| **Lived with both parents at GUp** |  |  |  |
| Not both parents | 961 (18.3%) | 421 (15.8%) | 540 (20.8%) |
| Both parents | 3955 (75.3%) | 2092 (78.6%) | 1863 (71.9%) |
| Missing | 337 (6.4%) | 149 (5.6%) | 188 (7.3%) |
| **Lived with both parents at AoW** |  |  |  |
| Not both parents | -- | 359 (13.5%) | -- |
| Both parents | -- | 1487 (55.9%) | -- |
| Missing | -- | 816 (30.6%) | -- |
| **Parental mental illness (birth to GUp)** |  |  |  |
| Yes | 2033 (38.7%) | 979 (36.8%) | 1054 (40.7%) |
| No/ Not recorded | 3220 (61.3%) | 1683 (63.2%) | 1537 (59.3%) |
| **Parental mental illness (birth to AoW)** |  |  |  |
| Yes | -- | 1001 (37.6%) | 1076 (41.5%) |
| No/ Not recorded | -- | 1661 (62.4%) | 1515 (58.5%) |
| **Any parental mental illness record (birth to GUp)** |  |  |  |
| Yes | 1557 (29.6%) | 738 (27.7%) | 819 (31.6%) |
| No/ Not recorded | 3696 (70.4%) | 1924 (72.3%) | 1772 (68.4%) |
| **Any parental mental illness record (birth to AoW)** |  |  |  |
| Yes | 1611 (30.7%) | 765 (28.7%) | 846 (32.7%) |
| No/ Not recorded | 3642 (69.3%) | 1897 (71.3%) | 1745 (67.3%) |
| **Any maternal mental illness self-reported (GUp)** |  |  |  |
| Possibly | 1102 (21.2%) | 539 (20.4%) | 563 (22.0%) |
| Possibly Not/ Not reported | 4099 (78.8%) | 2104 (79.6%) | 1995 (78.0%) |
| Missing | 52 (0.99%) | 19 (0.71%) | 33 (1.27%) |
| **Any paternal mental illness self-reported (GUp)** |  |  |  |
| Possibly | 35 (48.6%) | 13 (39.4%) | 22 (56.4%) |
| Possibly Not/ Not reported | 37 (51.4%) | 20 (60.6%) | 17 (43.6%) |
| Missing | 5181 (98.6%) | 2629 (98.8%) | 2552 (98.5%) |
| **Parental substance use (birth to GUp)** |  |  |  |
| Yes | 380 (7.23%) | 154 (5.8%) | 226 (8.7%) |
| No/ Not recorded | 4873 (92.8%) | 2508 (94.2%) | 2365 (91.3%) |
| **Parental substance use (birth to AoW)** |  |  |  |
| Yes | -- | 156 (5.9%) | 231 (8.9%) |
| No/ Not recorded | -- | 2506 (94.1%) | 2360 (91.1%) |
| **Any parental substance use record (birth to GUp)** |  |  |  |
| Yes | 87 (1.66%) | 41 (1.54%) | 46 (1.8%) |
| No/ Not recorded | 5166 (98.3%) | 2621 (98.5%) | 2545 (98.2%) |
| ***Any parental substance use record (birth to AoW)*** |  |  |  |
| Yes | 94 (1.79%) | 43 (1.6%) | 51 (2.0%) |
| No/ Not recorded | 5159 (98.2%) | 2619 (98.4%) | 2540 (98.0%) |
| ***Any parental hazardous alcohol use self-reported (GUp)*** |  |  |  |
| Yes | 312 (5.94%) | 123 (4.62%) | 189 (7.3%) |
| No | 4829 (91.9%) | 2506 (94.1%) | 2323 (89.7%) |
| Missing | 112 (2.1%) | 33 (1.24%) | 79 (3.0%) |
| **Child was bullied (birth to AoW)** |  |  |  |
| Yes | -- | 919 (34.5%) | -- |
| No | -- | 752 (28.2%) | -- |
| Missing | -- | 991 (37.2%) | -- |
| **Child was bullied (GUp)** |  |  |  |
| Yes | 1298 (24.7%) | 642 (24.1%) | 656 (25.3%) |
| No | 3853 (73.3%) | 1964 (73.8%) | 1889 (72.9%) |
| Missing | 102 (2.0%) | 56 (2.1%) | 46 (1.8%) |
| **Child was bullied (AoW)** |  |  |  |
| Yes | -- | 414 (15.6%) | -- |
| No | -- | 939 (35.3%) | -- |
| Missing | -- | 1309 (49.2%) | -- |
| Notes: * = missing data excluded; -- = not applicable; 1 = values presented are mean and standard deviation (SD) in brackets; 2 = latent class analysis classification [38] – the 5 classes are: Least dep = Least deprived and most educated, Employed not mat dep = Employed, not materially deprived, Employed no money = Employed, no access to money, Benefits but coping = Receiving benefits but not materially deprived, Most dep = Most economically deprived; *italic fonts* = components of the parental substance use or mental illness categories; AoW = at Age of Wonder Study (early adolescence, in 2022-23 and 2023-24 academic years); BiB = Born in Bradford Birth Cohort; birth = refers to the baseline survey shortly before childbirth; GUp = at Growing up Study (around mid-childhood); IMD = Index of Multiple Deprivation. |  |  |  |

## Results

### Overall sample characteristics

Among all 5253 eligible children, the mean age was 9.72 (SD 1.14) at GUp (mid-childhood), which also represents the average length of AE exposure between birth and mid-childhood. 64.5% of children were of Asian ethnicities, of which 84.7% were of Pakistani ethnicity; 28.7% were White, with 92.7% being White British; 4.1% were of mixed ethnicities; 1.2% were of Black ethnicities; and 1.4% were of Other ethnicities. 68.3% of children and their families could be classified as most deprived based on their address at baseline, according to the Index of Multiple Deprivation (IMD) quintile 2010 score. The largest proportion of children’s mothers received benefits but coping (i.e. not materially deprived) at baseline (26.9%), whereas the minority was considered to be most economically deprived (11.4%), classified with Fairley *et al.*’s [38] latent class analysis classification of maternal socioeconomic positions (SEPs).

For the 2662 eligible children included from baseline to early adolescence, the mean age at AoW (early adolescence) was 13.8 (SD 0.82). The proportions of each ethnic group and SEP at childbirth differed from those in the 5253 sample. The proportion of Asian children remained the largest and increased to nearly three-quarters (73.5%, of which 86.8% were of Pakistani ethnicity), while the proportions of White, Mixed, and Black ethnic groups decreased. Children of mothers who received benefits increased and remained the largest group (29.2%). The IMD 2010 score of this subset increased slightly (i.e. more deprived), which could be explained by the locations of participating schools in the AoW in the two included academic years (see Table 3 Characteristics of the included children and families, stratified by surveysTable 3).

### Sample prevalence of adverse experiences in childhood

For the period from birth to mid-childhood, 3147 children (59.9%, 95% CI [58.6%, 61.2%], of 5253) were classified as having experienced at least one of the four AEs. The most common AE was parental mental illness (38.7%, 95% CI [37.4%, 40.0%], n=2033), followed by bullying (24.7%, 95% CI [23.5%, 25.9%], n=1298), not living with both parents at mid-childhood (18.3%, 95% CI [17.3%, 19.3%], n=961), not living with both parents around birth (10.2%, 95% CI [9.4%, 11.0%], n=536), and parental substance use was the least common (7.2%, 95% CI [6.5%, 7.9%], n=380) (see Figure 3 and S3 Table 3 in S3 Supporting Information).

**Figure 3.**
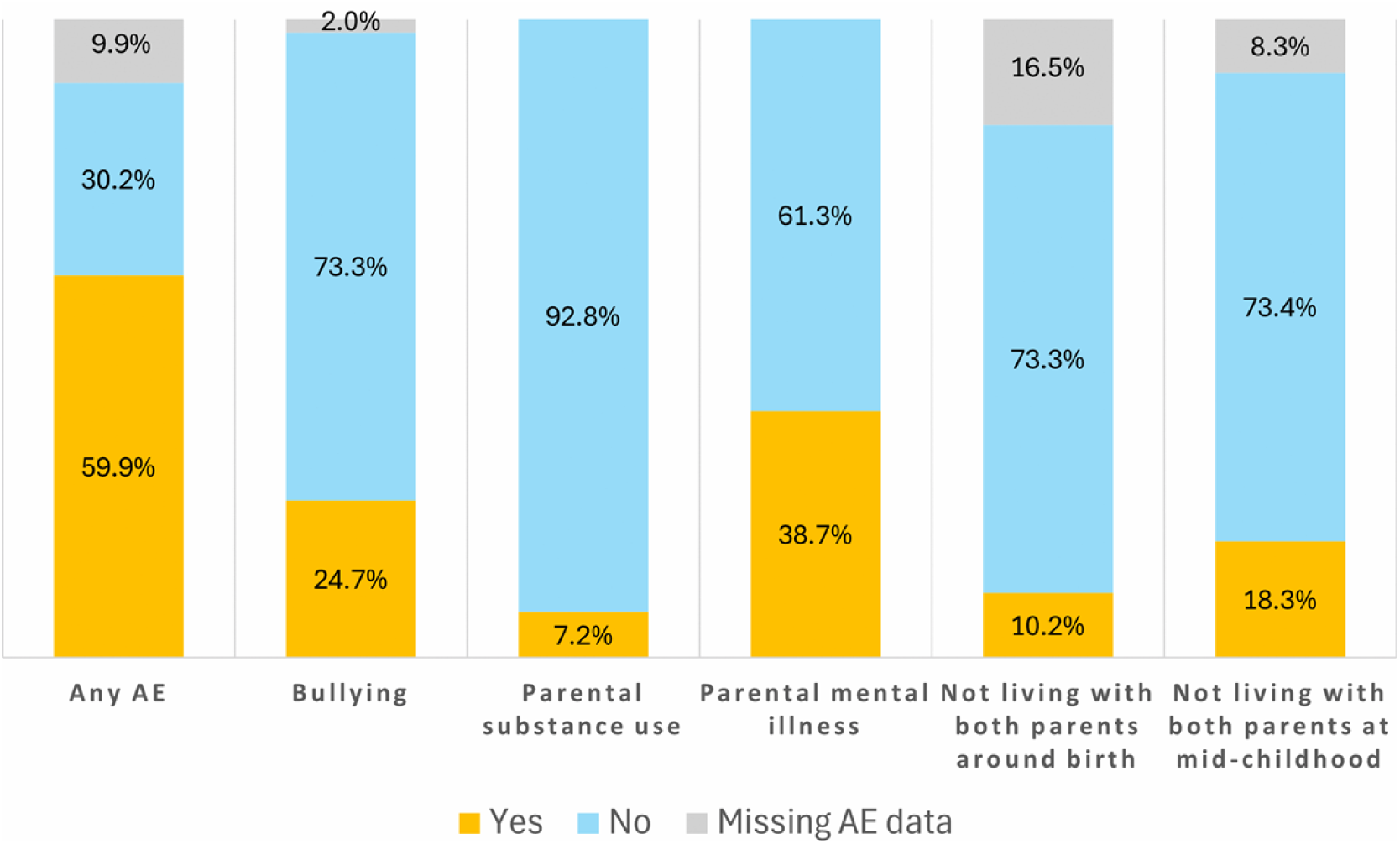
Percentages of children who experienced adverse experiences in childhood (AE) between birth and mid-childhood (n=5253)

For the birth-to-adolescence period, 1747 of 2662 (65.6%, 95% CI [63.8%, 67.4%]) eligible children were classified as having experienced at least one AE. The most common AE was parental mental illness (37.6%, 95% CI [35.8%, 39.4%], n=1001), followed by bullying (34.5%, 95% CI [32.7%, 36.3%], n=919), not living with both parents (25.6%, 95% CI [23.9%, 27.3%], n=681), and parental substance use (5.9%, 95% CI [5.0%, 6.8%], n=156) (see Figure 4 and S3 Table 4 in S3 Supporting Information).

**Figure 4.**
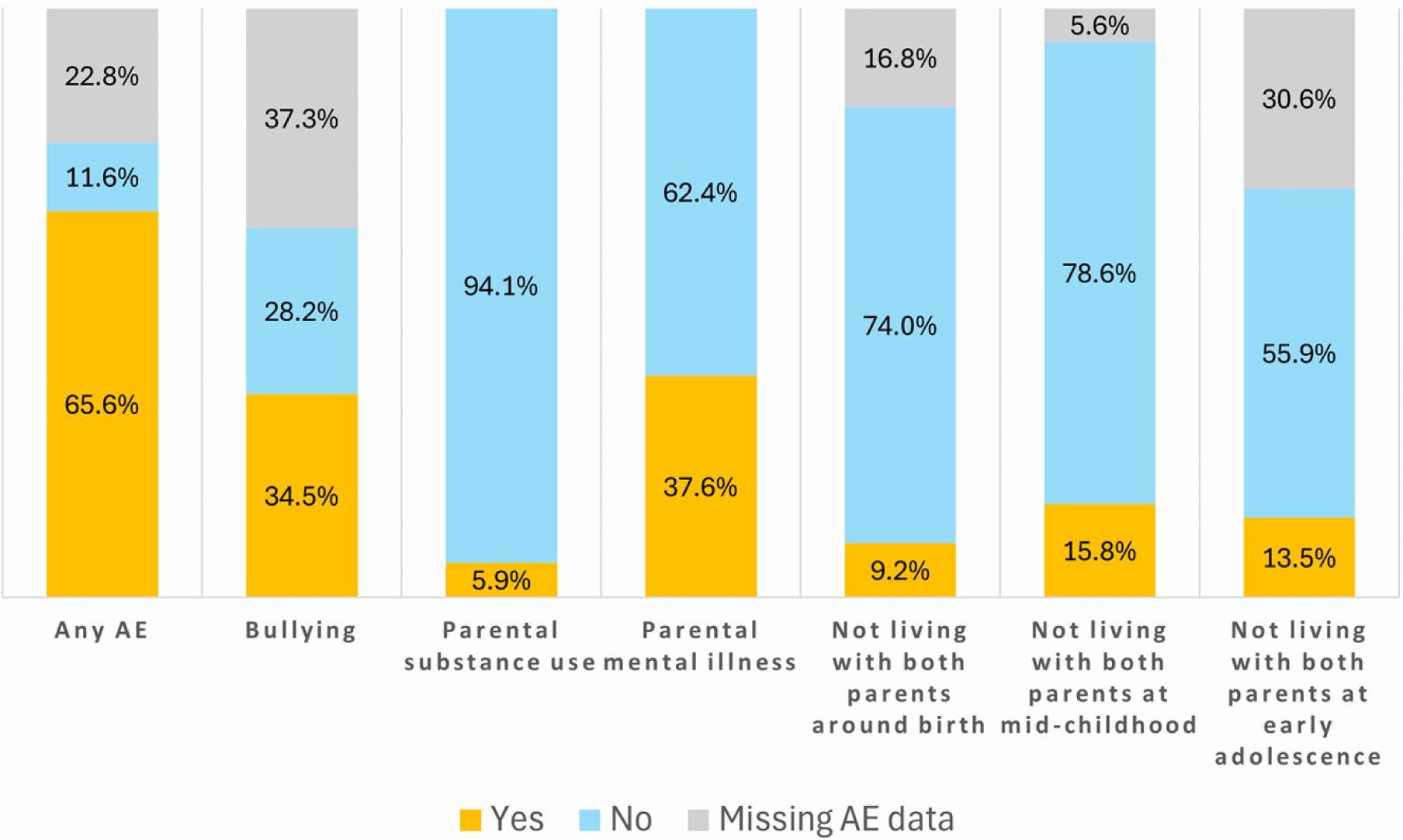
Percentages of children who experienced adverse experiences in childhood (AE) between birth and early adolescence (n=2662)

**Table 4.** Contingency table for number of mothers with mental illness according to GP records and self-report at baseline and GUp surveys (n=4651)

|  | GP record (Yes) | GP record (No) | Total |
| --- | --- | --- | --- |
| Self-report (Yes) | 571 | 410 | 981 |
| Self-report (No) | 811 | 2859 | 3670 |
| Total | 1377 | 3274 | 4651 |

The common patterns emerged are that children whose parents were relatively older at the time of childbirth (i.e. mean age over 28 for mothers and over 31 for fathers), whose mothers were least deprived and most educated or received benefits but coping, baseline household size was relatively bigger (i.e. mean of over 4.5), or whose ethnicity was Asian, were less likely to experience any or some of the four classified AEs compared to their peers. The proportions of children with any AE were similar between boys and girls, except for bullying.

### Comparison between information sources

After summarising data from various information sources for classification, we describe the differences observed in parental substance use and parental mental illness between parent-report surveys and GP records. The participants and measurements differed across the information sources used to classify these two AEs. Hence, the data were expected to differ between the sources. The findings from the surveys and healthcare records for classifying parental mental illness and parental substance use are detailed below, focusing on the sample of 5253 children from baseline (around birth) to 29 May 2024 (early adolescence).

4933 mothers and fathers of the 5253 children participated and self-reported any mental illness history and symptoms in GUp. 981 mothers and 28 fathers self-reported mental illness history and symptoms on the Patient Health Questionnaire (PHQ-8) [27] and/or Generalized Anxiety Disorder 7- item (GAD-7) questionnaire [28] questionnaires – a total of 1009 parents of 1135 children (21.6% of 5253). Table 4 shows that, notably, fewer mothers self-reported mental illness (n=981) than those with a GP record (n=1377). Only 41.5% (n=571) of mothers with a GP record also self-reported a history of mental illness. Additionally, none of the 1398 fathers who consented to data linkage, including the 28 who self-reported a history and symptoms, had a GP record of mental illness.

There were also different proportions of parents classified as having substance use across the three information sources. Table 5 and Table 6 show that there were more mothers and fathers who self-reported substance use than those whose substance use was recorded by a GP.

**Table 5.**
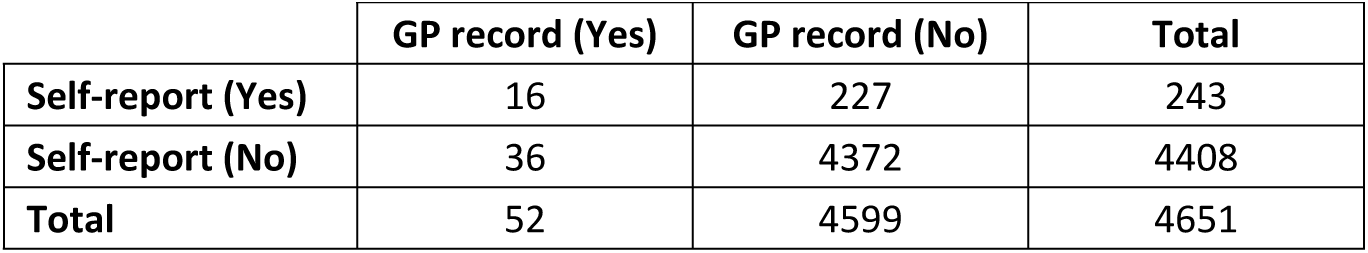
Contingency table for number of mothers with substance use according to GP records and self-report at baseline and GUp surveys (n=4651)

|  | GP record (Yes) | GP record (No) | Total |
| --- | --- | --- | --- |
| Self-report (Yes) | 16 | 227 | 243 |
| Self-report (No) | 36 | 4372 | 4408 |
| Total | 52 | 4599 | 4651 |

**Table 6.** Contingency table for number of fathers with substance use according to GP records and self-report at GUp survey (n=1398)

|  | GP record (Yes) | GP record (No) | Total |
| --- | --- | --- | --- |
| **Self-report (Yes)** | 3 | 48 | 51 |
| **Self-report (No)** | 37 | 1310 | 1347 |
| **Total** | 40 | 1358 | 1398 |

## Discussion

### Summary of findings

Approximately six in ten included BiB children have experienced at least one of the four classified AEs between birth and early adolescence. The most common AE was parental mental illness, while the least common was parental substance use. This overall sample prevalence aligns with findings from AE studies of other UK birth cohorts that directly measured AEs during childhood in surveys, such as the Avon Longitudinal Study of Parents and Children (ALSPAC) (e.g., reported by Russell *et al.* [31]) and the Millennium Cohort Study (MCS) (e.g., reported by Straatmann *et al.* [32]). Nonetheless, the findings in this study may be only indirectly comparable due to differences in measurement and data collection methods.

It is feasible to classify and identify four common AEs – bullying, parental substance use, parental mental illness, and not living with both parents – using the BiB datasets through a combination of surveys and routinely collected data. However, other widely recognised AEs, e.g., child maltreatment, cannot be identified or inferred from the BiB survey data.

The most common sociodemographic characteristics observed among children with AE(s) were White or Mixed ethnicities, whose mothers were most deprived or were employed with no access to money, smaller household size at birth, and younger parents. Nevertheless, the sociodemographic patterns varied across the four AEs. White children, or those whose mothers were most deprived, were more likely to experience bullying and parental mental illness. Mixed and Black children, or those whose mothers were most deprived, were less likely to live with both parents throughout childhood. Mixed and White children, or those whose mothers were employed but not materially deprived, were more likely to experience parental substance use.

The level of agreement between information sources varied for the sample prevalence of parental substance use and parental mental illness. Another observation was that the proportion of missing data for classifying each AE category was generally higher among children who experienced more than one AE, suggesting that some missing data could be related to the measured AEs. The proportion of missing data for AE classification also varied across time points and information sources. Therefore, it is beneficial to access and use multiple sources to classify AEs and to collate a more comprehensive and reliable prevalence estimate.

### Strengths

Our study was the first to conduct a thorough investigation of longitudinal Born in Bradford survey data and GP records to identify four AEs, spanning birth to adolescence. The strengths of this study’s methods for classifying AEs include using data collected at multiple time points, employing multiple measurement methods and multiple informants, and tracking as many BiB children from birth as possible across all included time points. The data are prospective and longitudinal, referring to either the moment of reporting or a relatively short period before reporting, e.g., two months for early adolescence bullying experiences.

Whenever possible, we used multiple information sources and/or variables to cross-check or supplement the data from each source, thereby increasing confidence in the reliability and validity of the multiple measurement methods [39, 40]. The differences in findings across the information sources indicate that using multiple sources to supplement one another is beneficial. Some plausible cases could have been missed if only GP records or survey data were used for these classifications. Although the information in these sources was not specifically designed to collect AE data, the questions were included alongside other topics in the BiB surveys, which might unintentionally lessen the discomfort in disclosing the information [41].

Another strength was that the temporality of each classified AE was specified to indicate the shortest period or the nearest time point when the AEs occurred, e.g., birth to mid-childhood or at mid-childhood. This approach helped avoid the generalisation that an AE happened “during childhood”, which typically refers to the first 18 years of a person’s life in ACE measures and literature, but undermines recognition of the varying impact of AEs occurring at distinct developmental stages throughout this long period [42]. We also explicitly detailed the variables and information sources used, the R code for constructing the AEs, and the rationale and assumptions underlying the classification methods. All of these details were documented in the pre-registered analysis plan and are reusable.

### Limitations

Despite the strengths and feasibility of this study’s methods, there were challenges throughout the process and limitations to the findings. First, a general limitation relates to the substantial amount of missing data, particularly in the AoW surveys and the smaller representation of Black, Mixed, and Other ethnic groups. These limited the generalisability of this study’s findings to these ethnic groups in the broader BiB cohort or in the Bradford District. Second, the frequency or severity of the AEs could not be established either.

Two additional limitations are potential misclassifications and difficulties in accurately inferring the timing or duration of AE occurrence.

### Reasons for possible misclassifications

Only a minority of eligible children had both parents participating in the BiB surveys. Therefore, it was uncertain whether both parents or a parent’s partner living with the child had any substance use or mental illness that could affect the child. Even for children with data from both parents, underreporting of parental substance use and parental mental illness could not be ruled out. This was evidenced by a comparison of parents’ self-reported information in surveys with GP records. Moreover, if participating parents did not report, this could be due to intentional non-reporting (i.e. a type of missing data) or the absence of the event. These reasons could not be discerned from the available survey data, which was a limitation.

Another potential misclassification concerned the selected CTV3 diagnostic codes. Although the CTV3 code has been used in UK NHS primary care record systems, there is neither a consistent nor an official source providing descriptions or translations from other codes, e.g., Read v2, ICD-10, or SNOMED codes, nor a complete list of codes for indicating a condition. Including codes for specific prescriptions related to mental illness and substance use may improve the sensitivity of the searches, but the specificity may decrease.

Misclassification of survey data may result from the sensitivity and specificity of the survey questions used to identify AEs. These questions aimed to gather factual information from parents; however, their reliability for classifying AE has not been assessed. For instance, the recommended alcohol use assessment tool and guidance are not designed for identifying parental substance use that is problematic to the person and their families. Instead, they aim to identify alcohol consumption or habits that increase or pose a higher risk to an individual’s health. Nonetheless, using parents’ reported alcohol consumption aligns with the questions used in other cohort studies, such as ALSPAC [43] and MCS [32], which classified drinking frequency of daily or 5-6 times per week as parental substance use, without counting the alcohol unit consumed. These questions may also highlight some signs of underlying reasons for occasional heavy or frequent drinking.

### Challenges in establishing timing and duration

The start and duration of the AEs experienced by the children could not be precisely determined in this study. Most of the selected questions did not focus on when or how long the AE-related events started, for example, when a child first experienced bullying. Additionally, only three time points were included over 12-15 years of the children’s childhood, so the gaps between them could not accurately reflect the duration of any AE.

Although the date of the first record of substance use and mental illness was available in the GP records, this information was unlikely to be the date when the parent first experienced those problems. Therefore, the timing and presence of such records depended on whether and when the parent sought consultation and the accuracy of the record. The duration of these two AEs could not be determined because the GP may not record when or whether these problems were resolved.

### Variables in BiB datasets considered but not used

Several sets of BiB survey questions were considered but not used to classify as AEs. They included alcohol consumption during pregnancy, General Health Questionnaire-28 (GHQ-28), status and quality of the parents’ relationship, parenting and support from parents and family for the child, and adverse neighbourhood or victimisation. The main reasons for exclusion were either low response rates, insufficient details to infer AE, or the categories not meeting the AE criteria set for this study. These variables are summarised in Appendix 7 in S3 Supporting Information and listed in the S2 Supporting Information.

Nevertheless, these variables may be relevant to practitioners, commissioners, and other researchers for similar classifications or as explanatory factors to understand the relationship between AEs and their impacts on children and families. Other routine records now linked to BiB, including health visiting [23], and children’s social care available via Connect Bradford data linkage [44], may contain information about other AEs. Examples of AEs recorded in the health and social care records include parental substance use, parental mental illnesses, parental separation, domestic abuse, child abuse, and neglect, which can be found in the maternity, GP, hospital stay, and health visiting records of the child and/or parents [45–47].

### Moving forward

The methodology for classifying the four AEs emphasises the importance of using multiple information sources to enrich AE category classification. It is crucial to clearly document the study’s aims and methods for AE classification to ensure transparency and reproducibility. Sharing these details with other researchers and practitioners on recording, collecting, and using AE data can highlight similarities and differences in study findings, which may help identify the causal pathways of AEs and their subsequent health and life conditions.

It is essential to continue collecting information from parents and children on AEs in the Born in Bradford studies’ surveys, including BiB and the Born in Bradford Better Start birth cohort, to inform local and national public health authorities and support more effective prevention strategies. Additionally, co-production and discussions with children and parents about feasible survey questions help to understand and address challenges in obtaining responses for some less preferred measures [48].

## Conclusion

Adverse experiences in childhood are common among children from birth to early adolescence. Given the participants’ multiethnic composition, the findings extend the AE literature by showing that children across ethnic groups and socioeconomic positions experienced these AEs, though prevalence varied notably by subgroup. Accessible interventions and appropriate family policies targeting the early years are crucial for supporting families and children at higher risk in meeting their needs, such as practical support for financial security, accessible childcare, and child-rearing strategies. Prioritising these structural supports, guided by continuously collected AE data, is critical to reducing AE prevalence and breaking the cycle of their lifelong negative impacts.

## Data Availability

The variables used are provided in the supporting information. The accompanying code is available at GitHub via https://github.com/NatalieYELam/adverse_experiences-/blob/main/AE-classification_Rcodes.html

## Acknowledgements

We are grateful to all the BiB participants, health professionals, schools, and researchers who have contributed their time, data, and enthusiasm to BiB. We thank Dr Dan Lewer for his suggestions and for supervising NL on the methodology and the writing of this report. We also thank Sarah Blower and Josie Dickerson for their support and suggestions throughout this study.

## Author Contributions

Natalie Lam: conceptualisation, formal analysis, investigation, methodology, software, project administration, visualisation, writing – original draft preparation, writing – review & editing Ruth Wadman: conceptualisation, methodology, supervision, writing – review & editing Aidan Watmuff: data curation, methodology, software, writing – review & editing Simon Gilbody: supervision, writing – review & editing

## Supplementary Materials

S1: STROBE checklist

S2: Variables and CTV3 codes used

S3 Supporting Information-Appendix

## References

1. Bellis MA, Wood S, Hughes K, Quigg Z, Butler N. Tackling adverse childhood experiences (ACES): state of the art and options for action. Cardiff: Public Health Wales. 2023. [Accessed 5 June 2023]. Available from: https://phwwhocc.co.uk/wp-content/uploads/2023/03/2023-01-state-of-the-art-report-eng.pdf.

2. Stevens AJ. Health Needs Assessment of Adverse Childhood Experiences in Bradford. City of Bradford Metropolitan District Council. 2019. [Accessed 23 August 2024]. Available from: https://jsna.bradford.gov.uk/media/wfbjghiv/adverse-childhood-experiences-2019.pdf.

3. PHE. No child left behind: A public health informed approach to improving outcomes for vulnerable children. London: Public Health England,; 2020; Accessed 23 August 2024]. Available from: https://assets.publishing.service.gov.uk/government/uploads/system/uploads/attachment_data/file/913764/Public_health_approach_to_vulnerability_in_childhood.pdf.

4. Hetherington K. Ending childhood adversity: a public health approach. Edinburgh: Public Health Scotland. 2020. [Accessed 26 January 2023]. Available from: https://publichealthscotland.scot/media/3096/ending-childhood-adversity-a-public-health-approach.pdf.

5. Welsh Government. Review of Adverse Childhood Experiences (ACE) policy: report. How the ACE policy has performed and how it can be developed in the future.: Welsh Government Health and Social Service; 2021; Accessed 3 March 2023]. Available from: https://www.gov.wales/review-adverse-childhood-experiences-ace-policy-report-html.

6. Asmussen K, Fischer F, Drayton E, McBride T. Adverse childhood experiences: What we know, what we don’t know, and what should happen next—Early Intervention Foundation. Early Intervention Foundation Report. 2020. [Accessed 6 April 2023]. Available from: https://www.eif.org.uk/report/adverse-childhood-experiences-what-we-know-what-we-dont-know-and-what-should-happen-next.

7. Bradford Council. Bradford District Children and Young People’s Strategy 2023-2025 City of Bradford Metropolitan District Council; 2023 [Accessed 7 January 2025]. Available from: https://www.bradford.gov.uk/children-young-people-and-families/reports-policies-projects-and-strategies/bradford-district-children-and-young-people-s-strategy/.

8. ONS. Estimates of the population for England and Wales: Mid-2022: 2023 local authority boundaires edition of this dataset. In: Statistics OfN, editor. UK. 2024. [Accessed 9 January 2025]. Available from: https://www.ons.gov.uk/file?uri=/peoplepopulationandcommunity/populationandmigration/populationestimates/datasets/estimatesofthepopulationforenglandandwales/mid20222023localauthorityboundaires/mye22tablesew2023geogs.xlsx.

9. DWP. Official Statistics: Children in low income families: local area statistics, financial year ending 2024. Department for Work & Pensions. 2025. [Accessed 1 May 2026]. Available from: https://www.gov.uk/government/statistics/children-in-low-income-families-local-area-statistics-2014-to-2024/children-in-low-income-families-local-area-statistics-financial-year-ending-2024#at-a-local-authority-level.

10. Fagan AA, Novak A. Adverse childhood experiences and adolescent delinquency in a high-risk sample: A comparison of White and Black youth. Youth Violence and Juvenile Justice. 2018;16(4):395–417. doi: 10.1177/1541204017735568.

11. Morrow AS, Villodas MT, Cunius MK. Prospective risk and protective factors for juvenile arrest among youth at risk for maltreatment. Child Maltreatment. 2019;24(3):286–98. doi: 10.1177/1077559519828819.

12. von Elm E, Altman DG, Egger M, Pocock SJ, Gøtzsche PC, Vandenbroucke JP. The Strengthening the Reporting of Observational Studies in Epidemiology (STROBE) statement: guidelines for reporting observational studies. The Lancet. 2007;370(9596):1453–7. doi: 10.1016/S0140-6736(07)61602-X.

13. Lam N, Wadman R, Lewer D, Gilbody S, Watmuff A. Understanding adverse experiences in childhood: the role and impact on subsequent adolescent mental health and well-being – a longitudinal study [Pre-registration]. 2024. doi: 10.17605/OSF.IO/YU5VT.

14. Kalmakis KA, Chandler GE. Adverse childhood experiences: towards a clear conceptual meaning. Journal of Advanced Nursing. 2013;70(7):1489–501. doi: 10.1111/jan.12329.

15. Kelly-Irving M, Lepage B, Dedieu D, Bartley M, Blane D, Grosclaude P, et al. Adverse childhood experiences and premature all-cause mortality. European Journal of Epidemiology. 2013;28(9):721–34. doi: 10.1007/s10654-013-9832-9.

16. Dube SR, Anda RF, Felitti VJ, Edwards VJ, Williamson DF. Exposure to Abuse, Neglect, and Household Dysfunction Among Adults Who Witnessed Intimate Partner Violence as Children: Implications for Health and Social Services. Violence Vict. 2002;(1):3–17. doi: 10.1891/vivi.17.1.3.33635.

17. Felitti VJMD, Facp, Anda RFMD, Ms, Nordenberg DMD, Williamson DFMS, et al. Relationship of Childhood Abuse and Household Dysfunction to Many of the Leading Causes of Death in Adults: The Adverse Childhood Experiences (ACE) Study. American Journal of Preventive Medicine. 1998;14(4):245–58. doi: 10.1016/S0749-3797(98)00017-8.

18. Lam N, Fairweather S, Lewer D, Prescott M, Undugoda P, Dickerson J, et al. The association between adverse childhood experiences and mental health, behaviour, and educational performance in adolescence: A systematic scoping review. PLOS Mental Health. 2024;1(5):e0000165. doi: 10.1371/journal.pmen.0000165.

19. BiB. Guidance and FAQs: Expression of Interest (EoI) form (v8 dated 11 November 2022). 2022. [Accessed 11 December 2023]. Available from: https://borninbradford.nhs.uk/our-data/guidance-and-faqs/.

20. Raynor P, BiB Collaborative Group. Born in Bradford, a cohort study of babies born in Bradford, and their parents: Protocol for the recruitment phase. BMC Public Health. 2008;8(1):327. doi: 10.1186/1471-2458-8-327.

21. Wright J, Small N, Raynor P, Tuffnell D, Bhopal R, Cameron N, et al. Cohort Profile: The Born in Bradford multi-ethnic family cohort study. International Journal of Epidemiology. 2013;42(4):978– 91. doi: 10.1093/ije/dys112.

22. McEachan RRC, Santorelli G, Watmuff A, Mason D, Barber SE, Bingham DD, et al. Cohort Profile Update: Born in Bradford. International Journal of Epidemiology. 2024:dyae037. doi: 10.1093/ije/dyae037.

23. Santorelli G, Mason D, Watmuff A, Badrick E, Hough A, Yang T, et al. Born in Bradford Data Summary v1.0: A Researcher’s Guide to the Born in Bradford Research Programme Data. 2025. [Accessed 19 March 2026]. Available from: https://borninbradford.nhs.uk/wp-content/uploads/2025/08/BiB_Data_Summary_v1.0_06August2025.pdf.

24. NHS. View your GP health record 2023 [updated 8 November 2023; Accessed 23 June 2025]. Available from: https://www.nhs.uk/nhs-services/gps/view-your-gp-health-record/.

25. WHO. International Statistical Classification of Diseases and Related Health Problems 10th Revision (ICD-10). 2019. [Accessed 1 May 2024]. Available from: https://icd.who.int/browse10/2019/en#/V.

26. Prady SL, Pickett KE, Petherick ES, Gilbody S, Croudace T, Mason D, et al. Evaluation of ethnic disparities in detection of depression and anxiety in primary care during the maternal period: combined analysis of routine and cohort data. Br J Psychiatry. 2016;208(5):453–61. Epub 2016/01/23. doi: 10.1192/bjp.bp.114.158832. PubMed PMID: 26795424; PubMed Central PMCID: PMCPMC4853643.

27. Kroenke K, Strine TW, Spitzer RL, Williams JB, Berry JT, Mokdad AH. The PHQ-8 as a measure of current depression in the general population. J Affect Disord. 2009;114(1-3):163–73. Epub 20080827. doi: 10.1016/j.jad.2008.06.026. PubMed PMID: 18752852.

28. Spitzer RL, Kroenke K, Williams JB, Löwe B. A brief measure for assessing generalized anxiety disorder: the GAD-7. Arch Intern Med. 2006;166(10):1092–7. doi: 10.1001/archinte.166.10.1092. PubMed PMID: 16717171.

29. Finkelhor D, Shattuck A, Turner H, Hamby S. Improving the Adverse Childhood Experiences Study Scale. JAMA Pediatrics. 2013;167(1):70–5. doi: 10.1001/jamapediatrics.2013.420.

30. Goodman R. The Strengths and Difficulties Questionnaire: a research note. J Child Psychol Psychiatry. 1997;38(5):581–6. Epub 1997/07/01. doi: 10.1111/j.1469-7610.1997.tb01545.x. PubMed PMID: 9255702.

31. Russell AE, Heron J, Gunnell D, Ford T, Hemani G, Joinson C, et al. Pathways between early-life adversity and adolescent self-harm: the mediating role of inflammation in the Avon Longitudinal Study of Parents and Children. Journal of child psychology and psychiatry, and allied disciplines. 2019;60(10):1094–103. doi: 10.1111/jcpp.13100.

32. Straatmann VS, Lai E, Law C, Whitehead M, Str, berg-Larsen K, et al. How do early-life adverse childhood experiences mediate the relationship between childhood socioeconomic conditions and adolescent health outcomes in the UK? Journal of epidemiology and community health. 2020;74(11):969–75. doi: 10.1136/jech-2020-213817.

33. Heinzen E, Sinnwell J, Atkinson E, Gunderson T, Dougherty G. arsenal: An Arsenal of “R” Functions for Large-Scale Statistical Summaries. R package version 3.6.3. 2021. Available from: https://CRAN.R-project.org/package=arsenal. doi: 10.32614/CRAN.package.arsenal.

34. Subirana I, Sanz H, Vila J. Building Bivariate Tables: The compareGroups Package for R. Journal of Statistical Software. 2014;57(12):1 – 16. doi: 10.18637/jss.v057.i12.

35. Wickham H, François R, Henry L, Müller K, Vaughan D. dplyr: A Grammar of Data Manipulation. R package version 1.1.4. 2023. Available from: https://CRAN.R-project.org/package=dplyr. doi: 10.32614/CRAN.package.dplyr.

36. Wickham H, Averick M, Bryan J, Chang W, McGowan LDA, François R, et al. Welcome to the Tidyverse. Journal of open source software. 2019;4(43):1686. doi: 10.21105/joss.01686.

37. Shire K, Newsham A, Rahman A, Mason D, Ryan D, A. Lawlor D, et al. Conducting longitudinal cohort research in secondary schools: Insights from the Born in Bradford Age of Wonder study. Wellcome Open Research. 2025;10(27). doi: 10.12688/wellcomeopenres.23534.1.

38. Fairley L, Cabieses B, Small N, Petherick ES, Lawlor DA, Pickett KE, et al. Using latent class analysis to develop a model of the relationship between socioeconomic position and ethnicity: cross-sectional analyses from a multi-ethnic birth cohort study. BMC Public Health. 2014;14(1):835. doi: 10.1186/1471-2458-14-835.

39. Murray J, Farrington DP, Eisner MP. Drawing conclusions about causes from systematic reviews of risk factors: The Cambridge Quality Checklists. Journal of Experimental Criminology. 2009;5(1):1–23. doi: 10.1007/s11292-008-9066-0.

40. Jolliffe D, Murray J, Farrington D, Vannick C. Testing the Cambridge Quality Checklists on a review of disrupted families and crime. Criminal Behaviour and Mental Health. 2012;22(5):303–14. doi: 10.1002/cbm.1837.

41. Quigg Z, Wallis S, Butler N. Routine enquiry about adverse childhood experiences implementation pack pilot evaluation. Liverpool John Moores University. 2018. Available from: https://www.gov.uk/government/publications/routine-enquiry-about-adverse-childhood-experiences-implementation-pack-evaluation.

42. Pollmann A, Bates KE, Fuhrmann D. A framework for understanding adverse adolescent experiences. Nature Human Behaviour. 2025;9(3):450–63. doi: 10.1038/s41562-024-02098-x.

43. Farooq B, Russell AE, Howe LD, Herbert A, Smith ADAC, Fisher HL, et al. The relationship between type, timing and duration of exposure to adverse childhood experiences and adolescent self-harm and depression: findings from three UK prospective population-based cohorts. Journal of Child Psychology and Psychiatry. 2024;65:1369–87. doi: 10.1111/jcpp.13986.

44. Sohal K, Mason D, Birkinshaw J, West J, McEachan RRC, Elshehaly M, et al. Connected Bradford: a Whole System Data Linkage Accelerator. Wellcome Open Res. 2022;7:26. Epub 2022/12/07. doi: 10.12688/wellcomeopenres.17526.2. PubMed PMID: 36466951; PubMed Central PMCID: PMCPMC9682213.

45. Lowthian E, Anthony R, Evans A, Daniel R, Long S, Bandyopadhyay A, et al. Adverse childhood experiences and child mental health: an electronic birth cohort study. BMC Medicine. 2021;19(1):172. doi: 10.1186/s12916-021-02045-x.

46. Syed S, Gonzalez-Izquierdo A, Allister J, Feder G, Li L, Gilbert R. Identifying adverse childhood experiences with electronic health records of linked mothers and children in England: a multistage development and validation study. The Lancet Digital Health. 2022;4(7):e482–e96. doi: 10.1016/S2589-7500(22)00061-9.

47. Parkin K. Holding pieces of the same puzzle: using an understanding of the prevalence and impact of adverse childhood experiences (ACEs) to target mental health resources. 2024. doi: 10.17863/CAM.119444.

48. Ryan D, Nutting H, Parekh C, Crookes S, Southgate L, Caines K, et al. Ready, set, co(produce): a co-operative inquiry into co-producing research to explore adolescent health and wellbeing in the Born in Bradford Age of Wonder project. Res Involv Engagem. 2024;10(1):41. Epub 20240430. doi: 10.1186/s40900-024-00578-y. PubMed PMID: 38689373; PubMed Central PMCID: PMCPMC11060965.

49. OpenSAFELY. OpenCodelists n.d. [Accessed 15 April 2024]. Available from: https://www.opencodelists.org/.

50. BioPortal. Read Codes, Clinical Terms Version 3 (CTV3) 2024 [updated 31 January 2024; Accessed 23 May 2024]. 2023AB:[Clinical Terms Version 3 (CTV) (Read Codes) (Q199): National Health Service National Coding and Classification Centre]. Available from: https://bioportal.bioontology.org/ontologies/RCD.

51. OpenSAFELY. Depression CTV3 2020 [updated 9 July 2020; Accessed 15 April 2024]. Available from: https://www.opencodelists.org/codelist/opensafely/depression/2020-07-09/.

52. OpenSAFELY. Psychosis, schizophrenia + bipolar affective disease 2020 [updated 9 July 2020; Accessed 16 April 2024]. Available from: https://www.opencodelists.org/codelist/opensafely/psychosis-schizophrenia-bipolar-affective-disease/2020-07-09/.

53. Denaxas S, Liu G, Feng Q, Fatemifar G, Bastarache L, Kerchberger EV, et al. Mapping the Read2/CTV3 controlled clinical terminologies to Phecodes in UK Biobank primary care electronic health records: implementation and evaluation. AMIA Annu Symp Proc. 2021;2021:362–71. Epub 20220221. PubMed PMID: 35308936; PubMed Central PMCID: PMCPMC8861677.

54. Bradford D. Read codes v2 (CTV2) 2022 [updated 10 July 2023; Accessed 15 April 2024]. Available from: 10.17605/OSF.IO/83S7M.

55. OpenSAFELY. Hazardous alcohol drinking CTV3 2021 [updated 26 January 2021; Accessed 15 April 2024]. Available from: https://www.opencodelists.org/codelist/opensafely/hazardous-alcohol-drinking/6364474b/.

56. Iwagami M, Tomlinson LA. Clinical codelist - Alcohol status read codes [Internet] London: London School of Hygiene & Tropical Medicine; 2020 [Accessed 21 May 2024]. Available from: https://datacompass.lshtm.ac.uk/id/eprint/1691/.

57. Adesanya E, Cook S, Mansfield KE, Crellin E, Smeeth L, Herrett E. Clinical Code list - Read codes for substance abuse [Internet] London: London School of Hygiene & Tropical Medicine; 2021 [Accessed 21 May 2024]. Available from: https://datacompass.lshtm.ac.uk/id/eprint/2185/.

58. Forray A, Merry B, Lin H, Ruger JP, Yonkers KA. Perinatal substance use: a prospective evaluation of abstinence and relapse. Drug Alcohol Depend. 2015;150:147–55. Epub 20150303. doi: 10.1016/j.drugalcdep.2015.02.027. PubMed PMID: 25772437; PubMed Central PMCID: PMCPMC4387084.

59. Babor TF, Higgins-Biddle JC, Saunders JB, Monteiro MG. AUDIT : the Alcohol Use Disorders Identification Test : guidelines for use in primary health care. Second ed. Geneva: World Health Organization. 2001. [Accessed 23 June 2025]. Available from: https://www.who.int/publications/i/item/WHO-MSD-MSB-01.6a.

60. NICE. Alcohol-use disorders: diagnosis and management of physical complications (CG100) 2017 [Accessed 19 April 2024]. Available from: https://www.nice.org.uk/guidance/cg100.

61. OHID. Alcohol use disorders identification test consumption (AUDIT C). 2020; Accessed 30 April 2024]. Available from: https://assets.publishing.service.gov.uk/media/6357a7d7e90e0777a45a9caa/Alcohol-use-disorders-identification-test-for-consumption-AUDIT-C_for-print.pdf.

62. NICE. Alcohol - problem drinking 2023 [Accessed 19 April 2024]. Available from: https://cks.nice.org.uk/topics/alcohol-problem-drinking/.

63. BiB. BiB Data Dictionary. n.d. Available from: https://borninbradford.github.io/datadict/index.html.

64. Goldberg DP, Gater R, Sartorius N, Ustun TB, Piccinelli M, Gureje O, et al. The validity of two versions of the GHQ in the WHO study of mental illness in general health care. Psychol Med. 1997;27(1):191–7. doi: 10.1017/s0033291796004242. PubMed PMID: 9122299.

65. Amorim M, Soares S, Abrahamyan A, Severo M, Fraga S. Patterns of childhood adversity and health outcomes in early adolescence: Results from the Generation XXI cohort. Preventive Medicine. 2023;171:107500. doi: 10.1016/j.ypmed.2023.107500.

